# A Model of Supply-Chain Decisions for Resource Sharing with an Application to Ventilator Allocation to Combat COVID-19

**DOI:** 10.1101/2020.04.02.20051078

**Authors:** Sanjay Mehrotra, Hamed Rahimian, Masoud Barah, Fengqiao Luo, Karolina Schantz

## Abstract

We present a stochastic optimization model for allocating and sharing a critical resource in the case of a pandemic. The demand for different entities peaks at different times, and an initial inventory for a central agency is to be allocated. The entities (states) may share the critical resource with a different state under a risk-averse condition. The model is applied to study the allocation of ventilator inventory in the COVID-19 pandemic by FEMA to different US states. Findings suggest that if less than 60% of the ventilator inventory is available for non-COVID-19 patients, FEMA’s stockpile of 20,000 ventilators (as of 03/23/2020) would be nearly adequate to meet the projected needs in slightly above average demand scenarios. However, when more than 75% of the available ventilator inventory must be reserved for non-COVID-19 patients, various degrees of shortfall are expected. In a severe case, where the demand is concentrated in the top-most quartile of the forecast confidence interval and states are not willing to share their stockpile of ventilators, the total shortfall over the planning horizon (till 05/31/20) is about 232,000 ventilator days, with a peak shortfall of 17,200 ventilators on 04/19/2020. Results are also reported for a worst-case where the demand is at the upper limit of the 95% confidence interval.

## 1 Introduction

COVID-19 was first identified in Wuhan, China in December 2019 [20]. It has since become a global pandemic. As of 03/31/2020 the United States has overtaken China in the number of deaths due to the disease, with more than 3,900 deaths. US, Italy, and Spain have all surpassed the death toll in China. However, United States tops the list of all countries in the current number of confirmed COVID-19 cases exceeding 400,000 [5]. In Northern Italy, one of the global epicenters of the pandemic, COVID-19 completely overwhelmed the healthcare system, forcing doctors into impossible decisions about which patients to save. Physicians on the front lines have shared accounts of how they must now weigh factors like age, comorbidities and probability of surviving prolonged intubation when deciding which patients with respiratory failure will receive mechanical ventilation [23].

### 1.1 A Resource Constrained Environment

While approximately 80% of COVID-19 cases are mild, the most severe cases of COVID-19 can result in respiratory failure, with approximately 5% of patients requiring treatment in an intensive care unit (ICU) with mechanical ventilation [28]. Mechanical ventilation is used to save the lives of patients whose lungs are so damaged that they can no longer pump enough oxygen into the blood to sustain organ function. It provides more oxygen than can be delivered through a nasal cannula or face mask, allowing the patient’s lungs time to recover and fight off the infection. Physicians in Italy have indicated that critical COVID-19 patients often need to be intubated for a prolonged period of time (15-20 days) [23], further exacerbating ventilator scarcity.

Limiting the death toll within the US depends on the ability to allocate sufficient numbers of ventilators to hard hit areas of the country before infections peak and ensuring that the inventory does not run out. Harder hit states (such as New York, Michigan and Louisiana) are desperately trying to acquire additional ventilators in anticipation of significant shortages in the near future. Yet in the absence of a coordinated federal response, reports have emerged of states finding themselves forced to compete with each other in order to obtain ventilators from manufacturers [1]. According to New York’s Governer Cuomo, the state has ordered 17,000 ventilators at the cost of $25,000/ventilator, but is expected to receive only 2,500 over the next two weeks [3]. As of 03/31/2020, according to the US presidential news briefing, more than 8,100 ventilators have been allocated by FEMA around the nation. Of these, 400 ventilators have been allocated to Michigan, 300 to New Jersey, 150 to Louisiana, 50 to Connecticut, and 450 to Illinois, in addition to the 4,400 given to New York [8].

Going forward, the federal response to the COVID-19 pandemic will require centralized decision-making around how to equitably allocate, and reallocate, limited supplies of ventilators to states in need. Projections from the Institute for Health Metrics and Evaluation at the University of Washington, which assume that all states will institute strict social distancing practices and maintain them until after infections peak, show states will hit their peak demand at different time points throughout the months of April and May. Many states are predicted to experience a significant gap in ICU capacity, and similar, if not greater, gaps in ventilator capacity, with the time point at which needs will begin to exceed current capacity varying by state [25].

### 1.2 Our Contributions

In response to the above problem, this paper presents a model for allocation and possible reallocation of ventilators that are available in the national stockpile. Importantly, computational results from the model also provide estimates of the shortfall of ventilators in each state under different future demand scenarios.

This modeling framework can be used to develop master plans that will allocate part of the ventilator inventory here-and-now, while allocating and reallocating the available ventilators in the future. The modeling framework incorporates conditions under which part of the historically available ventilator inventory is used for non-COVID-19 patients, who also present themselves for treatment along with COVID-19 patients. Thus, only a fraction of the historical ventilator inventory is available to treat COVID-19 patients. The remaining demand needs are met by allocation and re-allocation of available ventilators from FEMA and availability of additional ventilators through planned production. FEMA is assumed as the central agency that coordinates state-to-state ventilator sharing. The availability of inventory from a state for re-allocation incorporates a certain risk-aversion parameter. We present results while performing a what-if analysis under realistically generated demand scenarios using available ventilator demand data and ventilator availability data for different US states. An online planning tool is also developed and made available for use at https://covid-19.iems.northwestern.edu [9].

### 1.3 Organization

This paper is organized as follows. A review of the related literature is provided in Section 2. We present our resource allocation planning model, and its re-formulation in Section 3. Section 4 presents our computational results under different mechanical ventilator demand scenarios for the COVID-19 pandemic in the US. In Section 5, we introduce our companion online COVID-19 ventilator allocation and sharing planning tool. We end the paper with some discussion and concluding remarks.

## 2 Literature Review

A medical resource allocation problem in a disaster is considered in [29]. Victims’ deteriorating health conditions are modeled as a Markov chain, and the resources are allocated to optimize the total expected health recovery rate and reduce the total waiting time. Certain illustrative examples in a queuing network setting are also given in [29]. The problem of scarce medical resource allocation after a natural disaster using a discrete event simulation approach is investigated in [14]. Specifically, the authors in [14] investigate four resource-rationing principles: first come-first served, random, most serious first, and least serious first. It is found that without ethical constraints, the least serious first principle exhibits the highest efficiency. However, a random selection provides a relatively fairer allocation of services and a better trade-off with ethical considerations. Resource allocation in an emergency department in a multi-objective and simulation-optimization framework is studied in [16]. Simulation and queuing models for bed allocation are studied in [27, 18].

The problem of determining the levels of contact tracing to control spread of infectious disease using a simulation approach to a social network model is considered in [11]. A linear programming model is used in investigating the allocation of HIV prevention funds across states [15]. This paper suggests that in the optimal allocation, the funds are not distributed in an equitable manner. A linear programming model to derive an optimal allocation of healthcare resources in developing countries is studied in [17]. Differential equation-based systems modeling approach is used in [10] to find a geographic and demographic dependent way of distributing pandemic influenza vaccines based on a case study of A/H1N1 pandemic.

In a more recent COVID-19-related study, the author [21] proposes a probability model to estimate the effectiveness of quarantine and isolation on controlling the spread of COVID-19. In the context of ventilator allocation, a conceptual framework for allocating ventilators in a public emergency is proposed in [30]. The problem of estimating mechanical ventilator demand in the United States during an influenza pandemic was considered in [22]. In a high severity pandemic scenario, a need of 35,000 to 60,500 additional ventilators to avert 178,000 to 308,000 deaths was estimated. Robust models for emergency staff deployment in the event of a flu pandemic were studied in [12]. Specifically, the authors focused on managing critical staff levels during such an event, with the goal of minimizing the impact of the pandemic. Effectiveness of the approach was demonstrated through experiments using realistic data.

A method for optimizing stockpiles of mechanical ventilators, which are critical for treating hospitalized influenza patients in respiratory failure, is introduced in [19]. In a case-study, mild, moderate, and severe pandemic conditions are considered for the state of Texas. Optimal allocations prioritize local over central storage, even though the latter can be deployed adaptively, on the basis of real-time needs. Similar to this paper, the model in [19] uses an expected shortfall of ventilators in the objective function, while also considering a second criteria of total cost of ventilator stockpiling. However, the model in [19] does not consider distribution of ventilators over time. In the case of COVID-19, the ventilator demand is expected to peak at different times in different states, as the demand for each state has different trajectories. Only forecasts are available on how the demand might evolve in the future.

In this paper, we assume that the planning horizon is finite, and for simplicity we assume that reallocation decisions will be made at discrete times (days). Under certain demand conditions, the ventilators may be in short supply to be able to meet the demand. Our model is formulated as a stochastic program, and for the purpose of this paper, we reformulate and solve the developed model in its extensive form. We refer the reader to [13, 26] for a general description of this topic.

## 3 A Model for Ventilator Allocation

In this section, we present a multi-period planning model to allocate ventilators to different regions, based on their needs, for the treatment of critical patients. We assume that the demand for ventilators at each planning period is stochastic. We further assume that there is a central agency that coordinates the ventilator (re)location decisions. The ventilators’ (re)location is executed at the beginning of each time period. Once these decisions are made and executed, the states can use their inventory to treat patients. Both the federal agency and the states have to decide whether to reserve their inventory in anticipation of future demand or share it with other entities.

Before presenting the formulation, we list the sets, parameters, and decision variables that are used in the model.

- Sets and indices
  – *𝒩* : states (regions), indexed by *n* ∈ *𝒩* := *{*1, …, |*𝒩* |*}*,
  – *𝒯* : planning periods, indexed by *t* ∈ *𝒯* := *{*1, …, |*𝒯* |*}*,
  – Ω: ventilators’ demand scenarios, indexed by *ω* ∈ Ω := *{*1, …, |Ω|*}*,
- Deterministic parameters
  – *Y*_*n*_: the initial inventory of ventilators in region *n* ∈ *𝒩* at time period *t* = 0,
  – *I*: the initial inventory of ventilators in the central agency at the beginning of time period *t* = 0,
  – *Q*_*t*_: the number of ventilators produced during the time period *t −* 1 that become available at the beginning of time period *t* ∈ *𝒯*, for *t ≥* 1,
  – *γ*_*n*_: the percentage of the initial inventory of ventilators in region *n* ∈ *𝒩* that cannot be used to care for the patients at the critical level,
  – *τ*_*n*_: the percentage of the initial inventory of ventilators in region *n* ∈ *𝒩* that the region is willing to share with other regions, among those that can be used to care for patients at the critical level,
  – *ρ*_*n*_: the risk-aversion of region *n* ∈ *𝒩* to send their idle ventilators to the central agency to be shared with other regions,
- Stochastic parameter
  – 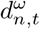: the number of patients in regions *n* ∈ *𝒩* at the critical level that need a ventilator at the beginning of time period *t* ∈ *𝒯* under scenario *ω* ∈ Ω,
  – **–** *p*^*ω*^: probability of scenario *ω* ∈ Ω,
- Decision variables
  – *x*_*n,t*_: the number of ventilators reallocated to region *n* ∈ *𝒩* by the central agency at the beginning of time period *t* ∈ *𝒯*,
  – 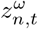: the number of ventilators reallocated to the central agency by region *n* ∈ *𝒯* at the beginning of time period *t* ∈ *𝒯* under scenario *ω* ∈ Ω,
  – 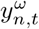: the number of ventilators at region *n* ∈ *𝒯* that can be used to care for the patients at the critical level at the end of time period *t* ∈ *{*0*} ∪ 𝒯* under scenario *ω* ∈ Ω,
  – 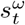: the number of ventilators at the central agency at the end of time period *t* ∈ *{*0*} ∪ 𝒯* under scenario *ω* ∈ Ω.

For notational convenience, we also define the vector 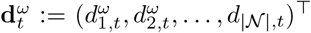 for *t* ∈ *𝒯* and *ω* ∈ Ω. Moreover, we define 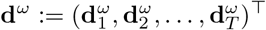, for *ω* ∈ Ω. We might drop the superscript *ω* ∈ Ω from this notation and use the same symbol with a tilde to denote that these parameters are stochastic. For example, we might use 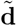. Similarly, we define the decision vector **x**_*t*_ := (*x*_1,*t*_, *x*_2,*t*_, …, *x*_|*𝒩*|,*t*_)^T^ for each *t* ∈ *𝒯* and **x** := (**x**_1_, **x**_2_, …, **x**_*T*_)^T^.

### 3.1 Model with No Lead-Time

In this section, we assume that there is no lead time between sending a ventilator by an entity (a region or the central agency) and delivery by another entity. With this assumption, the planning model to minimize the expected shortage of ventilators in order to treat patients at the critical level is formulated as a two-stage stochastic program as follows:

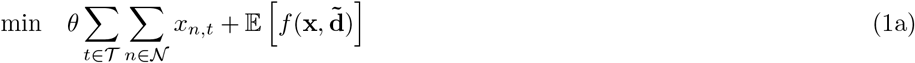

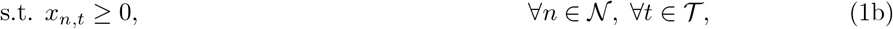

where

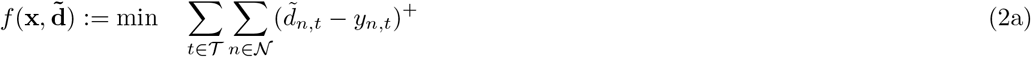

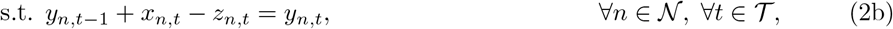

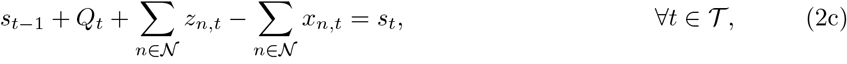

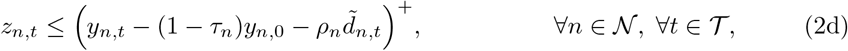

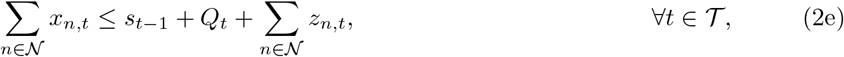

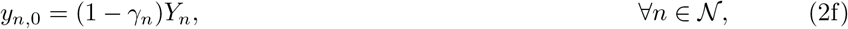

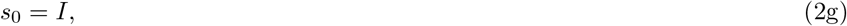

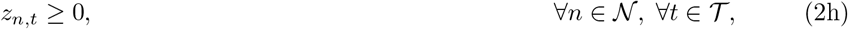

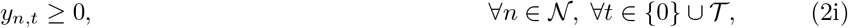

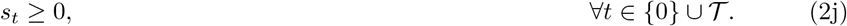

We now explain the model in detail. In the first stage, the central agency makes the “here-and-now” decisions **x** before the stochastic parameters 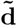 are realized. As captured in (1a), the goal of the central agency is to minimize the expected total shortage of ventilators over all time periods *t* ∈ *𝒯* and all regions *n* ∈ *𝒩*. The objective also includes a cost, parameterized by *θ* of allocating a ventilator by the central agency to a state at a given time. This cost can be set to zero, or set to a small value. In our computations we set *θ* = 0.01.

In the second stage, once the stochastic parameters 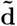 are realized, the “wait-and-see” decisions *z*_*n,t*_, *y*_*n,t*_, *s*_*t*_, *n* ∈ *𝒩* and *t* ∈ *𝒯*, are made. These decisions are scenario-specific, and are indicated by the superscript *ω* ∈ Ω, in the extensive formulation given in (3). Constraints (2b) and (2c) ensure the conservation of ventilators for the regions and the central agency at each time period, respectively. Constraint (2d) enforces that at each time period, a region is not sending out any ventilator to the central agency if its in-hand inventory is lower than its safety stock, where the safety stock is determined as 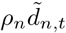, for *t* ∈ *𝒯* and *n* ∈ *𝒩*. Constraint (2e) ensures that at each time period, the total number of outgoing ventilators from the central agency to the regions cannot be larger than the available inventory, after incorporating the newly produced ventilators and the incoming ones from other regions. Constraints (2f) and (2g) set the initial inventory at the regions and central agency, respectively. The remaining constraints ensure the non-negativity of decision variables.

Note that the objective function (5a) and constraints (2d) are not linear. By introducing an additional variable, the term 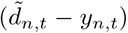^+^ in the objective function, for *n* ∈ *𝒩, t* ∈ *𝒯*, and *ω* ∈ Ω, can be linearized as

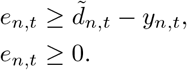

Furthermore, for each region *n* ∈ *𝒩* and time period *t* ∈ *𝒯*, constraint (2d) can be linearized as

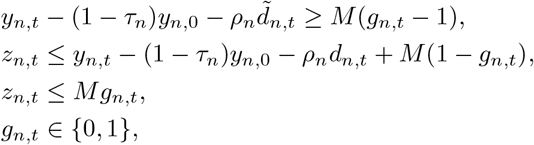

where *M* is a big number.

By incorporating the finiteness of the support of 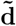, a linearized reformulation of model (1) can be written as a mixed-binary linear program in the following extensive form:

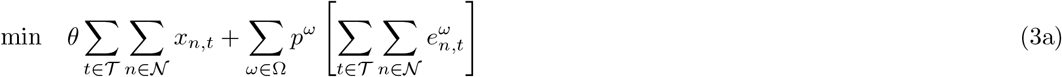

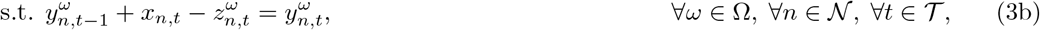

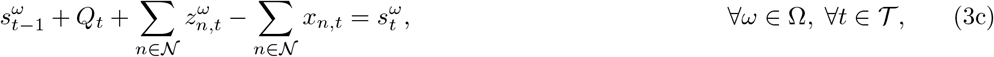

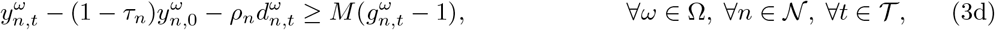

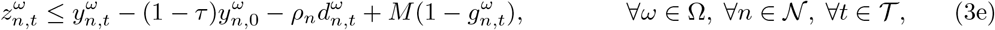

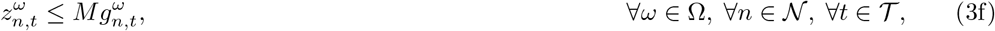

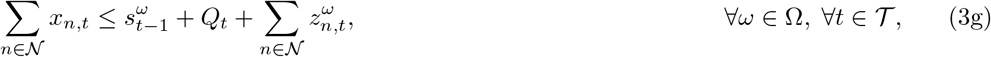

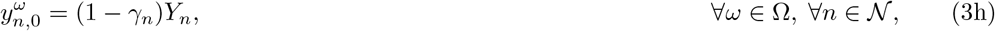

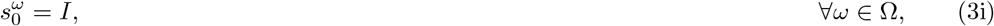

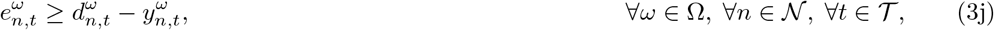

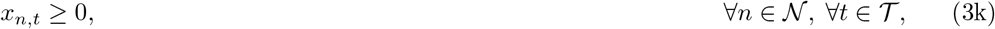

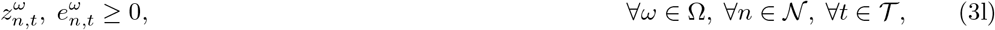

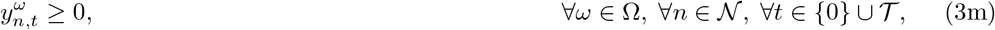

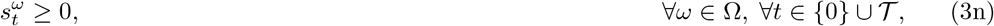

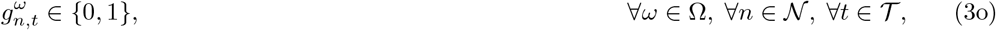

where 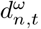 denotes the number of patients at the critical level in regions *n* ∈ *𝒩* that need a ventilator at the beginning of time period *t* ∈ *𝒯* under scenario *ω* ∈ Ω. Note that all second-stage variables 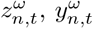, and 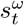, *n* ∈ *𝒩* and *t* ∈ *𝒯* in model (3) have superscript *ω* to indicate their dependence to scenario *ω* ∈ Ω. It is worth noting that (1) (and (3) as well) considers multi-period decisions. In the model, the central agency will make decisions for the entire planning horizon using the information that is available at the beginning of planning.

For our numerical experiments in Section 4, we used a commercial mixed-integer programming solver to obtain the results. Furthermore, we used *I* + *τ*_*n*_*y*_*n*,0_ + ∑ _*t*′≤*t*_ *Q*_*t*_ as a big-M for *n* ∈ *𝒩* and *t* ∈ *𝒯*.

### 3.2 Model with Lead-Time

In this section, we assume that there is a lead time of *L* time periods between sending a ventilator by an entity (a region or the central agency) and delivery by another entity. With this assumption, (1) can be generalized as follows:

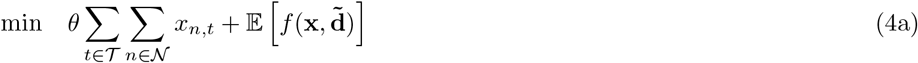

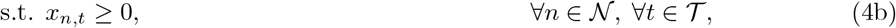

where

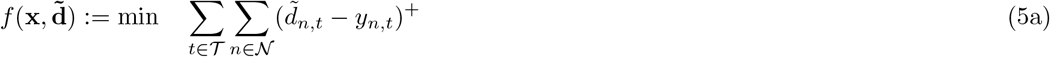

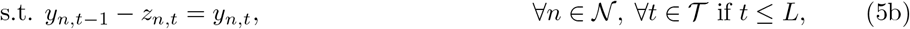

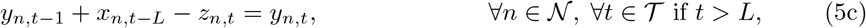

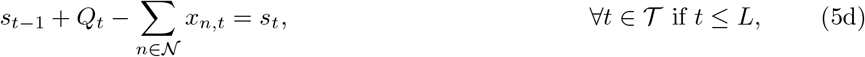

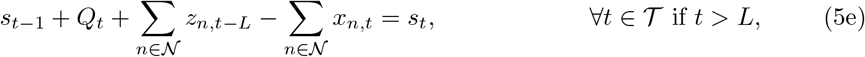

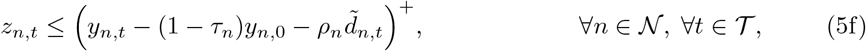

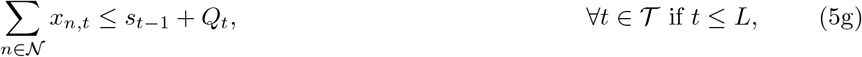

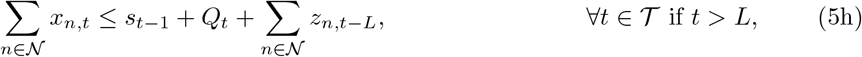

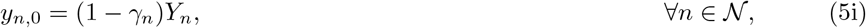

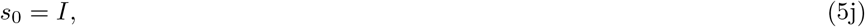

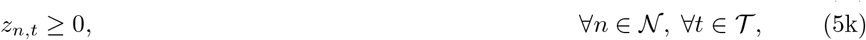

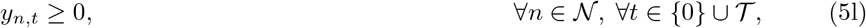

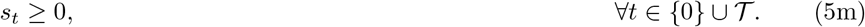

Note that by incorporating lead time in the planning model constraints (2b), (2c), and (2e) in model (2) have changed, and the remaining constraints are as in (2). Constraints (5b) and (5c) require the conservation of ventilators for the regions at each time period, where a ventilator sent by the federal agency to a region at time period *t −L, t > L*, will become available for the region at time period *t*. Constraints (5d) and (5e) ensure the conservation of ventilators for the central agency, respectively, where a ventilator sent by a region to the federal agency at time period *t −L, t > L*, will become available for the central agency at time period *t*. Constraint (5g) and (5h) enforce that the total number of outgoing ventilators from the central agency to the regions cannot be larger than the available inventory, after incorporating the newly produced ventilators and the incoming ones from other regions. Similar to (2b) and (2c), constraints (2e) are also divided into sets for *t* ≤*L* and *t > L* in (5g) and (5h).

By incorporating the finiteness of the support of 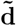, a linearized reformulation of model (1) can be written as a mixed-binary linear program in the following extensive form: Similar to (3), model (4) can be written as a mixed-binary linear program in the following extensive form:

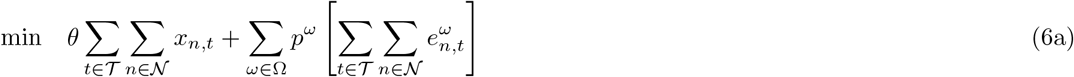

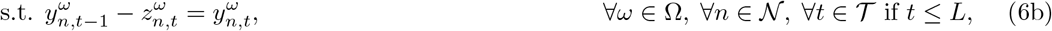

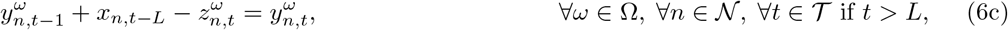

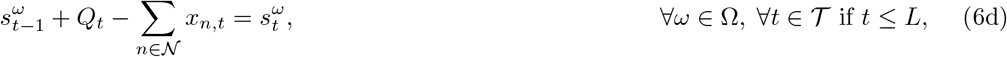

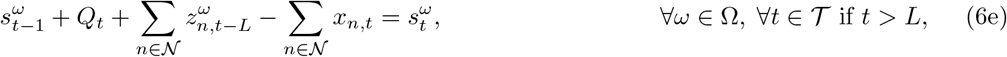

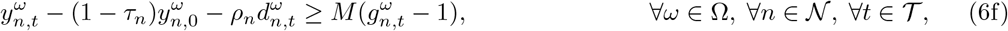

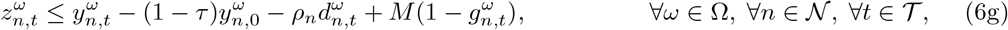

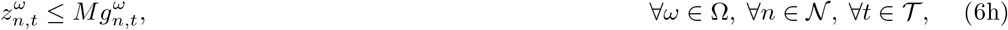

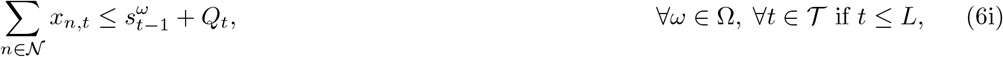

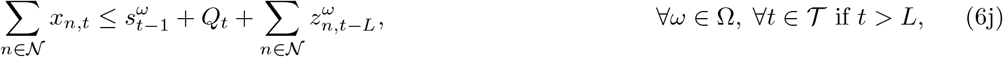

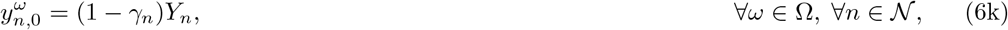

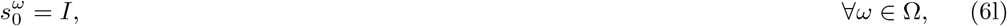

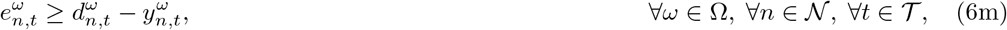

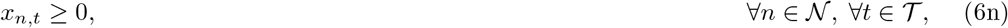

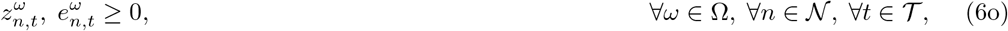

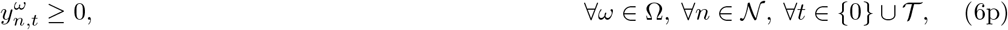

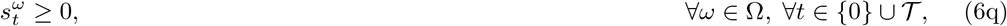

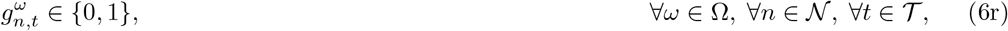

## 4 Ventilator Allocation Case Study: The US

The ventilator allocation model (3), described in Section 3, was implemented in Python 3.7. All computations were performed using GUROBI 9.0.1, on a Linux Ubuntu environment on two machines. In the first machine, we used 14 cores, with 3.4 GHz processor and 128 GB of RAM, and set the time limit to two hours. In the second machine, we used 64 cores, with 2.2 GHz processor and 128 GB of RAM, and set the time limit to three hours.

### 4.1 Ventilator Demand Data

Since projected ventilator need is a key input for the model, it is important to use accurate estimates of the demand forecasts. The forecasts of ventilator needs generated by [25] were used in our computational study. These forecasts were first made available on 03/26/2020, and used the most recent epidemiological data and advanced modeling techniques. The available information closely tracks the real-time data [4]. This COVID-19 needs forecast data was recently used in a recent presidential news brief [8]. Although it is difficult to validate the ventilator need forecasts against actual hospital and state level operational data, as this information is not readily available, we find that this model’s forecasts for deaths are quite accurate. For example, the model forecasted 217.9 deaths (CI: [176.95, 271.0]) on 03/29/2020 for NY state. The number of reported deaths in the state on 03/29/2020 were 237. Similarly, the model forecasted 262.2 deaths (CI: [206.9, 340]) on 03/30/2020 against the actual deaths of 253 on that day. A new update of the forecasts were published on 04/02/2020, and is used in the current version of our paper.

### 4.2 Demand Scenario Generation

We considered a seventy-day planning period, starting from 03/23/2020 and ending on 05/31/2020. We generated the random demands in ways that correspond to projected future demands under different mitigation effects. More precisely, we considered six different cases to generate random samples for the number of ventilators needed to care for COVID-19 patients. These cases are listed below:

Case I. **Average-I**: Each of the demand scenarios have equal probability and the distribution is uniform over the range of the CI provided in [25],

Case II. **Average-II**: The demand scenarios in the top 25% of CI have 0.25 probability (equally distributed); and scenarios in the bottom 75% have 0.75 probability,

Case III. **Above Average**: The demand scenarios in the top 25% of CI have 0.50 probability; and the scenarios in the bottom 75% have 0.50 probability.

Case IV. **Well Above Average**: The demand scenarios in the top 25% of CI have 0.75 probability; and the demand scenarios in the bottom 75% have 0.25 probability,

Case V. **Severe (CI Worst Quartile)**: The demand scenarios in top 25% have 100% probability, i.e., 0% probability is assigned for the bottom 75% demand scenarios.

Case VI. **Extreme (CI Upper Limit)**: There is only demand scenario, which happens at the upper limit of CIs.

We further discuss the demand generation procedure. A demand scenario contains the demand data for all days and states. In all Cases I–VI, we assumed that the forecast CI provided in [25], for each day and for each state, represents the support of the demand distribution.

Case I and II are generated to develop average demand scenario representations that use the information provided in the CI given in [25] in two different ways. In Case I, it is assumed that the mean is the median of the demand distribution (i.e., the right- and left-tail of the demand distribution have 0.5 probability). We randomly generated a number to indicate which tail to sample from, where both tails have the same 0.5 probability of being chosen. Once the tail is determined, we divided the tail into 50 equally-distanced partitions, and chose a random partition to uniformly sample from. We repeated this process for all days and states. We sampled from the same tail and partition for all days and states, although the range from which we sample depends on the CI. In this case, all scenarios are equally likely.

In Case II, we randomly generated a number to indicate which tail to sample from, where the top 25% of the CI (i.e., the right tail) has a 0.25 probability and the bottom 75% (i.e., the left tail) has a 0.75 probability of being chosen. If the right tail is chosen, we set the weight of the scenario to 0.25, and we set it to 0.75 otherwise. The rest of the procedure is similar to Case I. In order to determine the probability of scenarios, we normalized the weights. Demand scenarios in Cases III-V are generated in the same fashion as in Case II, where the only difference is in the probability of which tail to choose from, which is determined by the sampling scheme described in the definition of the case.

For Cases I-VI, we generated 24 scenarios, while in Case VI, there is only one scenario which happens at the upper limit of CI. Note that in each case, different quantities for the random demand 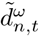 *t* ∈ *𝒯, n* ∈ *𝒩*, and *ω* ∈ Ω, might be generated. An illustration of the trajectory of demand scenarios over time is given in Figures 1–3 for the US and the States of New York and California.

**Figure 1:**
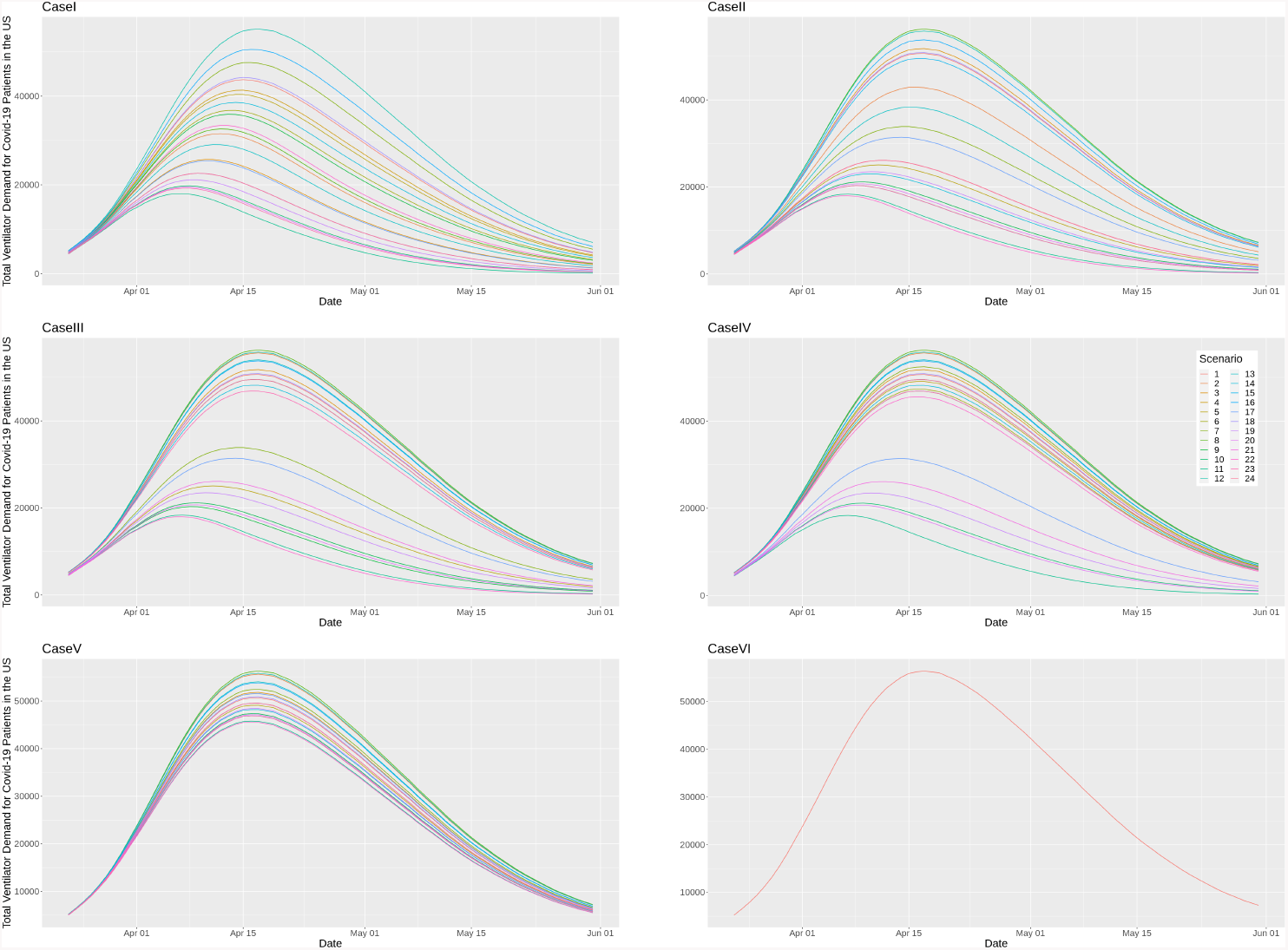
Summary of generated scenarios (Cases I–VI) for the US, adapted from the data provided in [25] on 04/02/2020.

**Figure 2:**
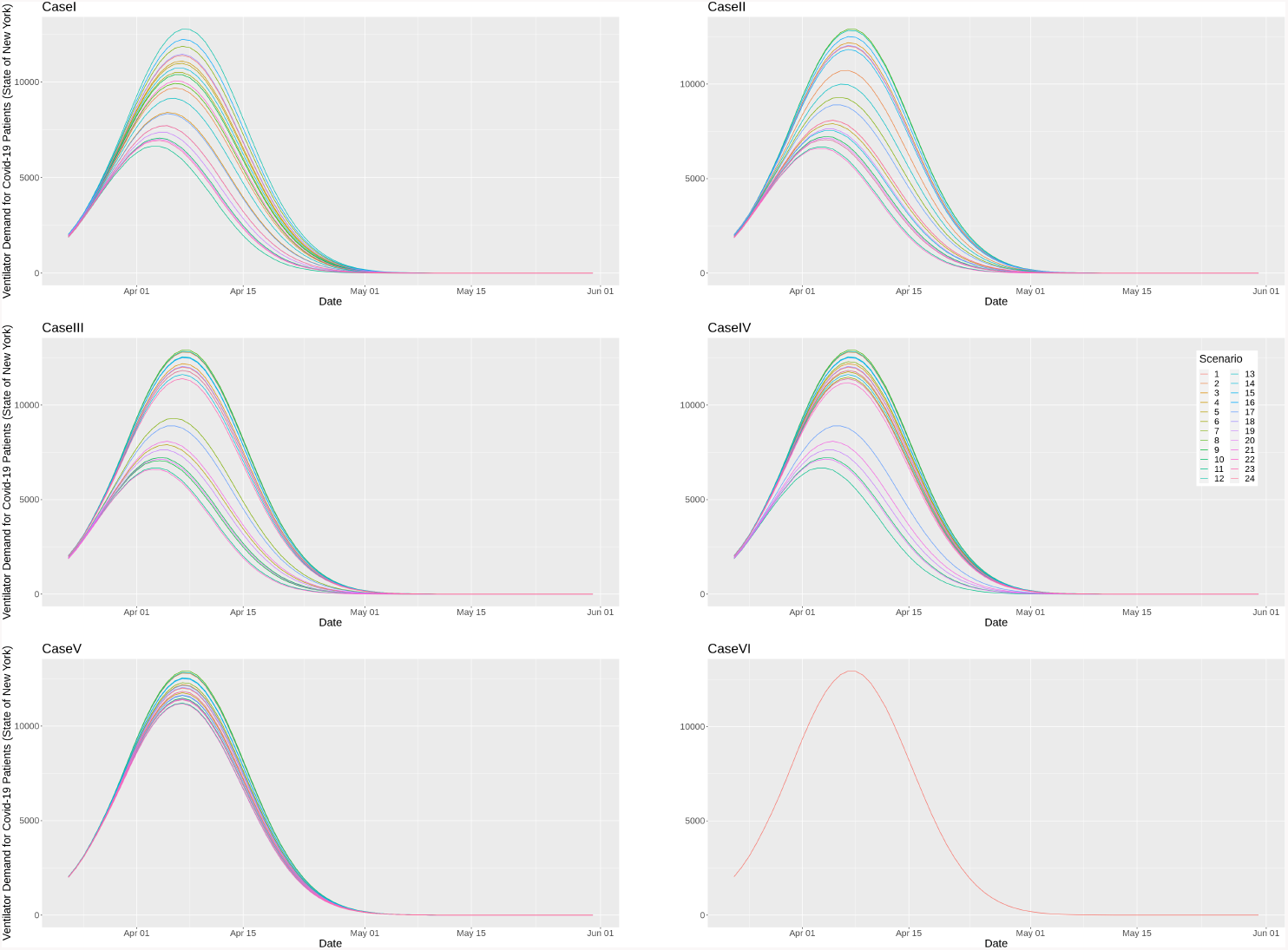
Summary of generated scenarios (Cases I–VI) for the State of New York, adapted from the data provided in [25] on 04/02/2020.

**Figure 3:**
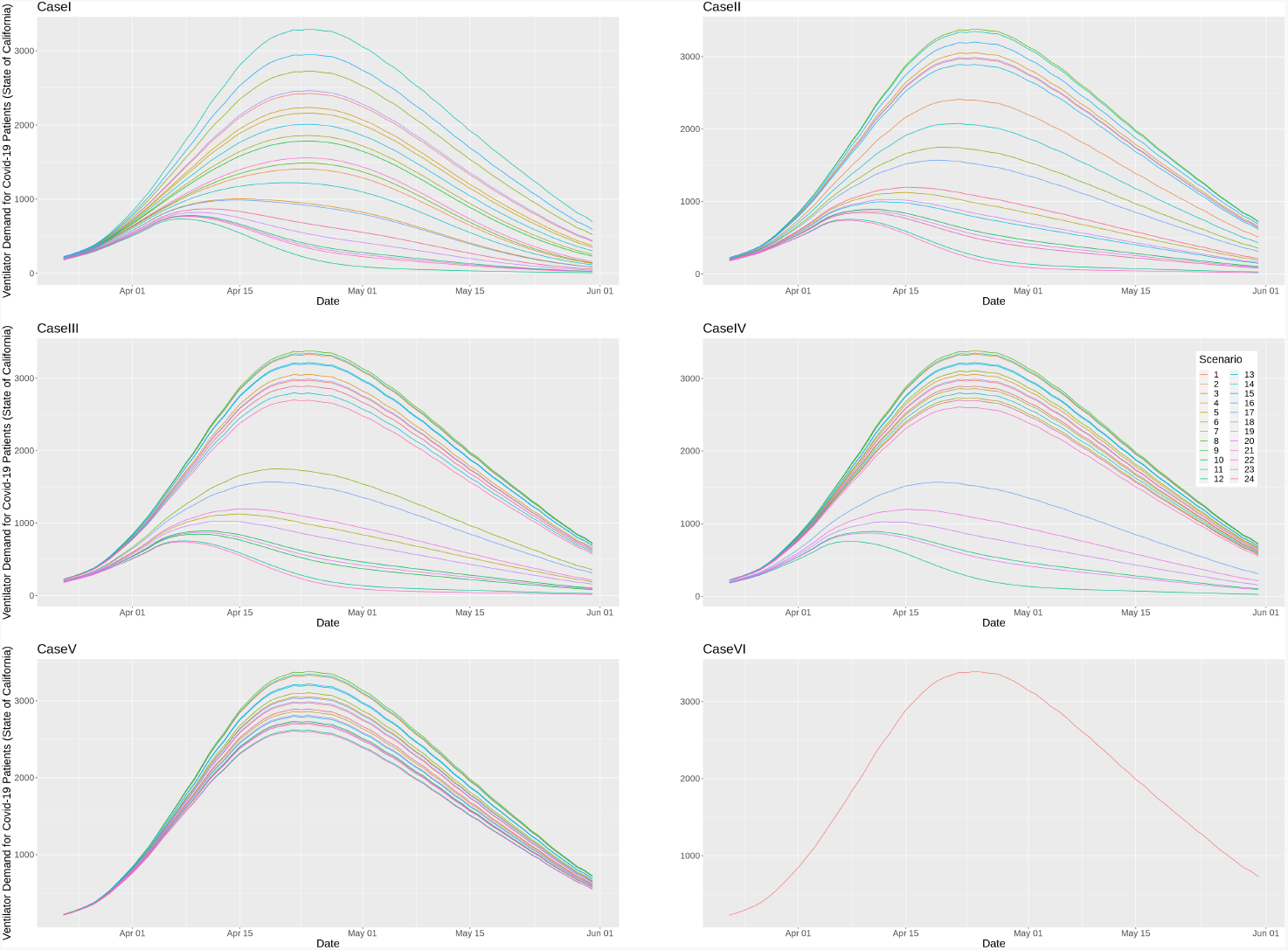
Summary of generated scenarios (Cases I–VI) for the State of California, adapted from the data provided in [25] on 04/02/2020.

### 4.3 Ventilator Inventory, Stockpile and Production

Another key input to the planning model is the initial ventilator inventory. As of 03/23/2020, before the rapid rise of COVID-19 cases in NY, FEMA had about 20,000 ventilators in reserve, i.e., *I* = 20, 000. We used this for our model which suggests ventilator allocation decisions from 03/23/2020.

Estimates for the initial inventory of ventilators at different states were obtained from [2]. These estimates are based on a hospital survey [24, 6]. The estimates for new ventilator production were obtained based on information provided at the US presidential briefings on 03/27/20 [7]. These estimates suggest that the normal yearly ventilator production capacity is about 30,000 ventilators/year. However, under the US Defense Production Act, with the participation of additional companies, production of approximately 10,000 ventilators/month could be possible [7]. Using this information, for the baseline case we assumed that the current daily ventilator production rate is *Q*_*t*_ = 80 ventilators/day; and it will be increased to *Q*_*t*_ = 320 ventilators/day starting on 04/15/2020. We refer to this case, as “baseline production”, and analyzed in Section 4.5.1. We also analyze the case that the ramp-up in production happens on 04/01/2020 or 04/07/2020 in Section 4.5.2.

### 4.4 Inventory Sharing Parameters

Recall that in the model, parameter *γ* is used to indicate the fraction of ventilators used to care for non-COVID-19 patients. Additionally, a parameter *τ* is used in the model to estimate a state’s willingness to share the fraction of their initial COVID-19-use ventilators. Similarly, the parameter *ρ* is used to control the state’s risk-aversion to sending their idle ventilators to FEMA for use in a different state. We suppose that for all states *n, n* ∈ *𝒩*, we have *γ*_*n*_ = *γ, ρ*_*n*_ = *ρ*, and *τ*_*n*_ = *τ*. In order to systematically study the ventilator allocations and shortfalls, we used the following parameters: *γ* ∈ *{*50%, 60%, 75%*}, ρ* ∈ *{*1.25, 1.5, 3*}*, and *τ* ∈ *{*0%, 10%, 25%*}*.

### 4.5 Numerical Results

In this section, we present and discuss the numerical results for the case that there is no lead-time, i.e., *L* = 0 or there is a lead-time of 1 day. For most instances, we observed that even obtaining an integer feasible solution to (3) and (4) in the time limit was not possible. Therefore, we replaced these models with their expected value problem, where the stochastic demand is substituted with the expected demand. Then, we solved the resulting model. This heuristic yields an integer feasible solution to model (3) and (3) for all instances we tested in the time limit, and we report those results here. In Section 4.5.1, we provide the results on ventilator’ shortage and inflow/outflow from/to FEMA for the case that there is no lead-time. We also analyze the effect of early ramp-up in production and lead-time on ventilators’ shortage in Sections 4.5.2 and 4.5.3, respectively.

#### 4.5.1 Baseline Production with No Lead-Time

In this section, for each setting (*γ, ρ, τ*), we solved the expected value problem of model (3) under Cases I–VI. A summary of ventilators’ shortage results is reported in Tables 1-3. We briefly describe the columns in these tables. Column “Total” denotes the expected total shortage, and is calculated as

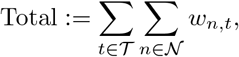

**Table 1:** Ventilators’ shortage summary under Cases I–VI, with *Q* = 320 and (*ρ, L*) = (1.25, 0).

|  |  |  |  |  |
| --- | --- | --- | --- | --- |
| (0.50, 0.00) | I | 3 | 0 | 0 |
|  | II | 1 | 0 | 0 |
|  | III | 4 | 0 | 0 |
|  | IV<sup>†</sup> | 3565 | 961 (04/15/2020) | 544 (04/16/2020, Illinois) |
|  | V<sup>†</sup> | 14966 | 2641 (04/16/2020) | 713 (04/16/2020, Alabama) |
|  | VI<sup>†</sup> | 121259 | 10513 (04/17/2020) | 2929 (04/16/2020, Alabama) |
| (0.50, 0.25) | I | 15 | 2 (04/21/2020) | 1 (04/22/2020, New Mexico) |
|  | II | 1 | 0 | 0 |
|  | III | 17 | 1 (04/24/2020) | 1 (04/07/2020, King and Snohomish Counties, WA) |
|  | IV<sup>†</sup> | 100 | 7 (04/17/2020) | 2 (04/21/2020, Idaho) |
|  | V<sup>†</sup> | 350 | 133 (04/16/2020) | 78 (04/16/2020, Alabama) |
|  | VI<sup>†</sup> | 73029 | 7512 (04/16/2020) | 2306 (04/16/2020, Alabama) |
| (0.50, 0.50) | I | 5 | 0 | 0 |
|  | II | 2 | 0 | 0 |
|  | III | 15 | 1 (03/28/2020) | 1 (04/20/2020, Other Counties, WA) |
|  | IV | 46 | 5 (04/15/2020) | 1 (04/24/2020, Idaho) |
|  | V<sup>†</sup> | 88 | 8 (04/19/2020) | 1 (04/25/2020, Montana) |
|  | VI<sup>†</sup> | 58379 | 6834 (04/17/2020) | 2283 (04/16/2020, Alabama) |
| (0.60, 0.00) | I | 6 | 1 (04/17/2020) | 0 |
|  | II | 4 | 0 | 0 |
|  | III | 27 | 3 (04/15/2020) | 1 (04/22/2020, Nebraska) |
|  | IV<sup>†</sup> | 29681 | 4317 (04/16/2020) | 1021 (04/16/2020, Alabama) |
|  | V<sup>†</sup> | 53698 | 6435 (04/16/2020) | 1608 (04/16/2020, Alabama) |
|  | VI<sup>†</sup> | 204905 | 15128 (04/17/2020) | 3827 (04/16/2020, Alabama) |
| (0.60, 0.25) | I | 10 | 1 (04/12/2020) | 1 (04/12/2020, Other Counties, WA) |
|  | II | 5 | 0 | 0 |
|  | III | 50 | 5 (04/19/2020) | 1 (04/19/2020, Rhode Island) |
|  | IV<sup>†</sup> | 14898 | 2726 (04/16/2020) | 757 (04/16/2020, Alabama) |
|  | V<sup>†</sup> | 34218 | 4548 (04/17/2020) | 1191 (04/16/2020, Alabama) |
|  | VI<sup>†</sup> | 163019 | 13334 (04/17/2020) | 3396 (04/16/2020, Alabama) |
| (0.60, 0.50) | I | 15 | 1 (04/26/2020) | 1 (03/25/2020, King and Snohomish Counties, WA) |
|  | II | 9 | 1 (04/05/2020) | 0 |
|  | III | 17 | 3 (04/19/2020) | 1 (04/21/2020, Iowa) |
|  | IV<sup>†</sup> | 8553 | 2031 (04/16/2020) | 600 (04/16/2020, Florida) |
|  | V<sup>†</sup> | 26916 | 3958 (04/17/2020) | 1110 (04/16/2020, Alabama) |
|  | VI<sup>†</sup> | 139404 | 12755 (04/16/2020) | 2835 (04/16/2020, Alabama) |
| (0.75, 0.00) | I | 10 | 1 (04/19/2020) | 0 |
|  | II | 23 | 2 (04/27/2020) | 1 (05/01/2020, South Dakota) |
|  | III<sup>†</sup> | 9390 | 1907 (04/15/2020) | 835 (03/23/2020, New York) |
|  | IV<sup>†</sup> | 131614 | 11430 (04/16/2020) | 2407 (04/16/2020, Alabama) |
|  | V<sup>†</sup> | 173004 | 13768 (04/16/2020) | 3001 (04/16/2020, Alabama) |
|  | VI<sup>†</sup> | 381943 | 22801 (04/17/2020) | 4411 (04/16/2020, Alabama) |
| (0.75, 0.25) | I | 56 | 6 (04/14/2020) | 1 (04/15/2020, Other Counties, WA) |
|  | II | 7 | 1 (04/22/2020) | 0 |
|  | III<sup>†</sup> | 5732 | 1350 (04/14/2020) | 835 (03/23/2020, New York) |
|  | IV<sup>†</sup> | 118583 | 10637 (04/16/2020) | 2395 (04/16/2020, Alabama) |
|  | V<sup>†</sup> | 157319 | 13337 (04/16/2020) | 2988 (04/16/2020, Alabama) |
|  | VI<sup>†</sup> | 359230 | 22200 (04/17/2020) | 4382 (04/16/2020, Alabama) |
| (0.75, 0.50) | I | 36 | 3 (04/14/2020) | 1 (04/14/2020, Maine) |
|  | II | 8 | 1 (05/01/2020) | 0 |
|  | III<sup>†</sup> | 3608 | 953 (04/14/2020) | 835 (03/23/2020, New York) |
|  | IV<sup>†</sup> | 109542 | 10446 (04/16/2020) | 2407 (04/16/2020, Alabama) |
|  | V<sup>†</sup> | 146505 | 12774 (04/15/2020) | 2837 (04/16/2020, Alabama) |
|  | VI<sup>†</sup> | 336767 | 21321 (04/17/2020) | 4345 (04/16/2020, Alabama) |

<sup>†</sup> Reached the time limit. The reported results correspond to the best integer solution found.

**Table 2:** Ventilators’ shortage summary under Cases I–VI, with *Q* = 320 and (*ρ, L*) = (1.50, 0).

| $(\gamma, \tau)$ | Case | Total | Worst date ($t$) | Worst date-state ($t, n$) |
| --- | --- | --- | --- | --- |
| (0.50, 0.00) | I | 5 | 0 | 0 |
|  | II | 1 | 0 | 0 |
|  | III | 3 | 0 | 0 |
|  | IV<sup>†</sup> | 6775 | 1677 (04/16/2020) | 459 (04/17/2020, Illinois) |
|  | V<sup>†</sup> | 20009 | 3352 (04/17/2020) | 713 (04/16/2020, Alabama) |
|  | VI<sup>†</sup> | 130989 | 11206 (04/17/2020) | 3164 (04/16/2020, Alabama) |
| (0.50, 0.25) | I | 4 | 0 | 0 |
|  | II | 1 | 0 | 0 |
|  | III | 3 | 0 | 0 |
|  | IV<sup>†</sup> | 30 | 4 (04/17/2020) | 1 (04/24/2020, Idaho) |
|  | V<sup>†</sup> | 4370 | 1105 (04/16/2020) | 352 (04/16/2020, Alabama) |
|  | VI<sup>†</sup> | 85197 | 8719 (04/17/2020) | 2461 (04/16/2020, Alabama) |
| (0.50, 0.50) | I | 3 | 0 | 0 |
|  | II | 4 | 0 | 0 |
|  | III | 8 | 1 (04/15/2020) | 0 |
|  | IV<sup>†</sup> | 47 | 4 (04/24/2020) | 1 (05/02/2020, Florida) |
|  | V<sup>†</sup> | 531 | 199 (04/17/2020) | 142 (04/17/2020, Florida) |
|  | VI<sup>†</sup> | 75157 | 7850 (04/17/2020) | 2359 (04/16/2020, Alabama) |
| (0.60, 0.00) | I | 4 | 0 | 0 |
|  | II | 4 | 0 | 0 |
|  | III | 10 | 2 (04/19/2020) | 1 (04/22/2020, Nebraska) |
|  | IV<sup>†</sup> | 37140 | 5209 (04/16/2020) | 1117 (04/16/2020, Alabama) |
|  | V<sup>†</sup> | 62710 | 7059 (04/17/2020) | 1644 (04/16/2020, Alabama) |
|  | VI<sup>†</sup> | 216449 | 15348 (04/18/2020) | 3449 (04/16/2020, Alabama) |
| (0.60, 0.25) | I | 8 | 1 (04/01/2020) | 0 |
|  | II | 4 | 0 | 0 |
|  | III | 36 | 4 (04/20/2020) | 1 (04/22/2020, Utah) |
|  | IV<sup>†</sup> | 23097 | 3827 (04/17/2020) | 756 (04/16/2020, Alabama) |
|  | V<sup>†</sup> | 46101 | 5944 (04/17/2020) | 1524 (04/16/2020, Alabama) |
|  | VI<sup>†</sup> | 178867 | 14269 (04/17/2020) | 3404 (04/16/2020, Alabama) |
| (0.60, 0.50) | I | 11 | 1 (04/28/2020) | 1 (04/30/2020, Hawaii) |
|  | II | 11 | 1 (04/19/2020) | 1 (04/19/2020, Idaho) |
|  | III | 29 | 4 (04/01/2020) | 1 (04/01/2020, New Jersey) |
|  | IV<sup>†</sup> | 16658 | 3001 (04/17/2020) | 862 (04/18/2020, Illinois) |
|  | V<sup>†</sup> | 36029 | 5012 (04/18/2020) | 1179 (04/16/2020, Alabama) |
|  | VI<sup>†</sup> | 157337 | 13400 (04/17/2020) | 3347 (04/16/2020, Alabama) |
| (0.75, 0.00) | I | 16 | 1 (04/16/2020) | 0 |
|  | II | 30 | 2 (04/22/2020) | 1 (04/04/2020, New Jersey) |
|  | III<sup>†</sup> | 14997 | 2790 (04/15/2020) | 835 (03/23/2020, New York) |
|  | IV<sup>†</sup> | 143039 | 12376 (04/17/2020) | 2674 (04/16/2020, Alabama) |
|  | V<sup>†</sup> | 185441 | 14537 (04/17/2020) | 3324 (04/16/2020, Alabama) |
|  | VI<sup>†</sup> | 394587 | 23457 (04/18/2020) | 4802 (04/16/2020, Alabama) |
| (0.75, 0.25) | I | 26 | 3 (04/14/2020) | 0 |
|  | II | 10 | 1 (05/01/2020) | 0 |
|  | III<sup>†</sup> | 11209 | 2388 (04/15/2020) | 835 (03/23/2020, New York) |
|  | IV<sup>†</sup> | 131086 | 11824 (04/16/2020) | 2674 (04/16/2020, Alabama) |
|  | V<sup>†</sup> | 172744 | 13957 (04/17/2020) | 2998 (04/16/2020, Alabama) |
|  | VI<sup>†</sup> | 375323 | 23105 (04/18/2020) | 4802 (04/16/2020, Alabama) |
| (0.75, 0.50) | I | 51 | 4 (04/14/2020) | 1 (04/15/2020, New Hampshire) |
|  | II | 13 | 1 (04/09/2020) | 1 (04/09/2020, Washington) |
|  | III<sup>†</sup> | 8698 | 2082 (04/15/2020) | 835 (03/23/2020, New York) |
|  | IV<sup>†</sup> | 122866 | 11314 (04/16/2020) | 2407 (04/16/2020, Alabama) |
|  | V<sup>†</sup> | 161075 | 13839 (04/17/2020) | 3001 (04/16/2020, Alabama) |
|  | VI<sup>†</sup> | 357178 | 22452 (04/18/2020) | 4384 (04/16/2020, Alabama) |

<sup>†</sup> Reached the time limit. The reported results correspond to the best integer solution found.

**Table 3:** Ventilators’ shortage summary under Cases I–VI, with *Q* = 320 and (*ρ, L*) = (3.00, 0).

| $(\gamma, \tau)$ | Case | Total | Worst date ($t$) | Worst date-state ($t, n$) |
| --- | --- | --- | --- | --- |
| (0.50, 0.00) | I | 3 | 0 | 0 |
|  | II | 1 | 0 | 0 |
|  | III | 3 | 0 | 0 |
|  | IV<sup>†</sup> | 22239 | 2974 (04/19/2020) | 917 (04/21/2020, Illinois) |
|  | V<sup>†</sup> | 48434 | 5591 (04/19/2020) | 1626 (04/21/2020, Florida) |
|  | VI<sup>†</sup> | 247524 | 18006 (04/17/2020) | 4387 (04/16/2020, Alabama) |
| (0.50, 0.25) | I | 3 | 0 | 0 |
|  | II | 1 | 0 | 0 |
|  | III | 3 | 0 | 0 |
|  | IV<sup>†</sup> | 7749 | 1250 (04/20/2020) | 410 (04/20/2020, Illinois) |
|  | V<sup>†</sup> | 25576 | 3348 (04/19/2020) | 1003 (04/16/2020, Alabama) |
|  | VI<sup>†</sup> | 190771 | 14883 (04/17/2020) | 3916 (04/16/2020, Alabama) |
| (0.50, 0.50) | I | 3 | 0 | 0 |
|  | II | 1 | 0 | 0 |
|  | III | 3 | 0 | 0 |
|  | IV<sup>†</sup> | 2666 | 750 (04/19/2020) | 289 (04/20/2020, Florida) |
|  | V<sup>†</sup> | 14403 | 2567 (04/20/2020) | 1115 (04/21/2020, Florida) |
|  | VI<sup>†</sup> | 132855 | 11965 (04/18/2020) | 3161 (04/21/2020, Illinois) |
| (0.60, 0.00) | I | 4 | 0 | 0 |
|  | II | 4 | 0 | 0 |
|  | III | 8 | 1 (04/24/2020) | 0 |
|  | IV<sup>†</sup> | 62677 | 6506 (04/19/2020) | 1529 (04/16/2020, Alabama) |
|  | V<sup>†</sup> | 100644 | 9489 (04/19/2020) | 2435 (04/23/2020, Illinois) |
|  | VI<sup>†</sup> | 337955 | 22532 (04/17/2020) | 4831 (04/16/2020, Alabama) |
| (0.60, 0.25) | I | 4 | 0 | 0 |
|  | II | 4 | 0 | 0 |
|  | III | 12 | 1 (04/20/2020) | 0 |
|  | IV<sup>†</sup> | 50770 | 5565 (04/19/2020) | 1271 (04/16/2020, Alabama) |
|  | V<sup>†</sup> | 81225 | 8120 (04/19/2020) | 1969 (04/16/2020, Alabama) |
|  | VI<sup>†</sup> | 297366 | 20457 (04/17/2020) | 4581 (04/16/2020, Alabama) |
| (0.60, 0.50) | I | 7 | 1 (03/24/2020) | 0 |
|  | II | 7 | 1 (04/17/2020) | 0 |
|  | III | 16 | 1 (04/19/2020) | 1 (04/06/2020, New York) |
|  | IV<sup>†</sup> | 39464 | 4727 (04/19/2020) | 1021 (04/16/2020, Alabama) |
|  | V<sup>†</sup> | 64578 | 7000 (04/19/2020) | 1618 (04/16/2020, Alabama) |
|  | VI<sup>†</sup> | 246365 | 17870 (04/18/2020) | 3954 (04/22/2020, Illinois) |
| (0.75, 0.00) | I | 7 | 1 (04/19/2020) | 0 |
|  | II | 9 | 1 (05/01/2020) | 0 |
|  | III<sup>†</sup> | 29710 | 3463 (04/19/2020) | 835 (03/23/2020, New York) |
|  | IV<sup>†</sup> | 179044 | 14176 (04/19/2020) | 2829 (04/16/2020, Alabama) |
|  | V<sup>†</sup> | 231612 | 17201 (04/19/2020) | 3429 (04/16/2020, Alabama) |
|  | VI<sup>†</sup> | 527275 | 30596 (04/17/2020) | 5700 (04/16/2020, Alabama) |
| (0.75, 0.25) | I | 23 | 3 (04/15/2020) | 1 (04/15/2020, Ohio) |
|  | II | 22 | 2 (04/08/2020) | 2 (04/09/2020, Washington) |
|  | III<sup>†</sup> | 27362 | 3410 (04/19/2020) | 835 (03/23/2020, New York) |
|  | IV<sup>†</sup> | 169753 | 13504 (04/19/2020) | 2674 (04/16/2020, Alabama) |
|  | V<sup>†</sup> | 218080 | 16487 (04/19/2020) | 3324 (04/16/2020, Alabama) |
|  | VI<sup>†</sup> | 506928 | 29776 (04/17/2020) | 5700 (04/16/2020, Alabama) |
| (0.75, 0.50) | I | 20 | 1 (04/14/2020) | 1 (04/22/2020, Indiana) |
|  | II | 8 | 1 (05/01/2020) | 0 |
|  | III<sup>†</sup> | 24204 | 3155 (04/19/2020) | 835 (03/23/2020, New York) |
|  | IV<sup>†</sup> | 160279 | 12675 (04/19/2020) | 2407 (04/16/2020, Alabama) |
|  | V<sup>†</sup> | 205062 | 15889 (04/19/2020) | 3289 (04/16/2020, Alabama) |
|  | VI<sup>†</sup> | 464863 | 28015 (04/17/2020) | 5447 (04/16/2020, Alabama) |

<sup>†</sup> Reached the time limit. The reported results correspond to the best integer solution found.

Where 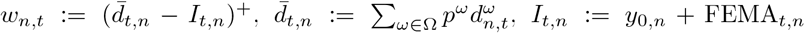, and 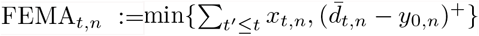.

Quantity “Worst day” in column “Worst day (*t*)” denotes the expected shortage in the worst day, and is calculated as

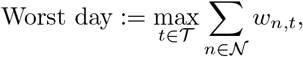

where *t* denotes a day that the worst expected shortage happens, i.e., *t* ∈ arg max_*t*∈*𝒯*_ ∑_∈ *𝒩*_ *w*_*n,t*_. Moreover, quantity “Worst day-state” in column “Worst day-state (*t*)” denotes the expected shortage in the worst day and state, and is calculated as

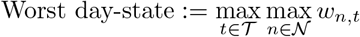

where (*t, n*) ∈ arg max_*t*∈*𝒯*_ arg max_*n*∈*𝒩*_ *w*_*n,t*_.

The results in Tables 1–3 suggest that when up to 60% of a state’s ventilator inventory is used for non-COVID-19 patients, FEMA’s current stockpile of 20,000 ventilators is nearly sufficient to meet the demand imposed by COVID-19 patients in mild cases (i.e., Cases I-III). The ventilator availability situation gets worse in the case where 75% (or greater %) of the available ventilators must be used for non-COVID-19 patients and states’ risk-aversion parameter to send the idle ventilators to FEMA to be used in a different state is 3. In this case, if states are willing to share up to 50% of their excess inventory with other states, then 12,700 number of ventilators will be required beyond FEMA’s current stockpile to meet demand in Cases I-IV. However, if no such sharing is considered, then the need for ventilators would increase to 14,200. This situation gets even worse for Cases V and VI, where the inventory shortfall on the worst day (04/19/2020) is between 17,200-30,600. This shortfall decreases moderately to 15,900-28,000 if states are willing to share part of their initial ventilator inventory. If parameter *ρ* goes down to 1.25, the inventory shortfall on the worst day (04/19/2020) is between 13,800-22,800. This shortfall decreases moderately to 12,800-21,300 if states are willing to share part of their initial ventilator inventory.

We also analyzed the ventilators’ reallocation to/from different states for the setting (*γ, ρ, τ*) = (0.75, 3, 0), which is the most dramatic case we considered from the inventory and stockpile perspectives. We report a summary of results in Table 4 under the two worst demand situations, Cases V (Severe) and VI (Extreme). Column “Total inflow” in this table denotes the total incoming ventilators to a state *n* ∈ *𝒩* from FEMA, and is calculated as

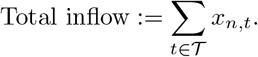

**Table 4:** Inflow and outflow from/to FEMA summary with *Q* = 320 and (*γ, ρ, τ, L*) =(0.75, 3.00, 0.00, 0).

| State | Case V |  |  | Case VI |  |  |
| --- | --- | --- | --- | --- | --- | --- |
|  | Total inflow | Total outflow | Net flow | Total inflow | Total outflow | Net flow |
| Alabama | 2025.00 | 0.00 | 2025.00 | 1496.00 | 0.00 | 1496.00 |
| Alaska | 53.00 | 0.00 | 53.00 | 50.00 | 0.00 | 50.00 |
| Arizona | 451.00 | 0.00 | 451.00 | 486.00 | 0.00 | 486.00 |
| Arkansas | 185.00 | 0.00 | 185.00 | 197.00 | 0.00 | 197.00 |
| California | 1154.00 | 0.00 | 1154.00 | 1445.00 | 0.00 | 1445.00 |
| Colorado | 762.00 | 0.00 | 762.00 | 702.00 | 0.00 | 702.00 |
| Connecticut | 242.00 | 0.00 | 242.00 | 183.00 | 0.00 | 183.00 |
| Delaware | 33.00 | 0.00 | 33.00 | 13.00 | 0.00 | 13.00 |
| District of Columbia | 0.00 | 0.00 | 0.00 | 0.00 | 0.00 | 0.00 |
| Florida | 4307.00 | 0.00 | 4307.00 | 5131.00 | 0.00 | 5131.00 |
| Georgia | 1057.00 | 0.00 | 1057.00 | 1051.00 | 0.00 | 1051.00 |
| Hawaii | 162.00 | 0.00 | 162.00 | 184.00 | 0.00 | 184.00 |
| Idaho | 208.00 | 0.00 | 208.00 | 202.00 | 0.00 | 202.00 |
| Illinois | 4361.00 | 53.50 | 4307.50 | 4555.00 | 0.00 | 4555.00 |
| Indiana | 910.00 | 0.00 | 910.00 | 1050.00 | 0.00 | 1050.00 |
| Iowa | 641.00 | 0.00 | 641.00 | 664.00 | 0.00 | 664.00 |
| Kansas | 282.00 | 0.00 | 282.00 | 304.00 | 0.00 | 304.00 |
| Kentucky | 166.00 | 0.00 | 166.00 | 231.00 | 0.00 | 231.00 |
| King and Snohomish Counties, WA | 212.00 | 0.00 | 212.00 | 209.00 | 0.00 | 209.00 |
| Louisiana | 615.00 | 4.75 | 610.25 | 543.00 | 230.00 | 313.00 |
| Maine | 105.00 | 0.00 | 105.00 | 92.00 | 0.00 | 92.00 |
| Maryland | 1324.00 | 0.00 | 1324.00 | 1511.00 | 0.00 | 1511.00 |
| Massachusetts | 589.00 | 0.00 | 589.00 | 447.00 | 0.00 | 447.00 |
| Michigan | 1354.00 | 42.08 | 1311.92 | 1037.00 | 885.00 | 152.00 |
| Minnesota | 289.00 | 0.00 | 289.00 | 254.00 | 0.00 | 254.00 |
| Mississippi | 434.00 | 0.00 | 434.00 | 443.00 | 0.00 | 443.00 |
| Missouri | 204.00 | 0.00 | 204.00 | 294.00 | 0.00 | 294.00 |
| Montana | 112.00 | 0.00 | 112.00 | 114.00 | 0.00 | 114.00 |
| Nebraska | 129.00 | 0.00 | 129.00 | 130.00 | 0.00 | 130.00 |
| Nevada | 332.00 | 0.00 | 332.00 | 337.00 | 0.00 | 337.00 |
| New Hampshire | 106.00 | 0.00 | 106.00 | 92.00 | 0.00 | 92.00 |
| New Jersey | 718.00 | 19.71 | 698.29 | 505.00 | 384.00 | 121.00 |
| New Mexico | 204.00 | 0.00 | 204.00 | 195.00 | 0.00 | 195.00 |
| New York | 11144.00 | 420.75 | 10723.25 | 14187.00 | 13544.00 | 643.00 |
| North Carolina | 656.00 | 0.29 | 655.71 | 727.00 | 0.00 | 727.00 |
| North Dakota | 35.00 | 0.00 | 35.00 | 33.00 | 0.00 | 33.00 |
| Ohio | 216.00 | 0.00 | 216.00 | 176.00 | 0.00 | 176.00 |
| Oklahoma | 325.00 | 0.00 | 325.00 | 274.00 | 0.00 | 274.00 |
| Oregon | 114.00 | 0.00 | 114.00 | 149.00 | 0.00 | 149.00 |
| Other Counties, WA | 181.00 | 0.00 | 181.00 | 193.00 | 0.00 | 193.00 |
| Pennsylvania | 209.00 | 0.00 | 209.00 | 76.00 | 0.00 | 76.00 |
| Rhode Island | 68.00 | 0.00 | 68.00 | 64.00 | 0.00 | 64.00 |
| South Carolina | 324.00 | 0.00 | 324.00 | 353.00 | 0.00 | 353.00 |
| South Dakota | 81.00 | 0.00 | 81.00 | 93.00 | 0.00 | 93.00 |
| Tennessee | 866.00 | 0.00 | 866.00 | 590.00 | 0.00 | 590.00 |
| Texas | 2471.00 | 0.00 | 2471.00 | 3010.00 | 0.00 | 3010.00 |
| Utah | 209.00 | 0.00 | 209.00 | 227.00 | 0.00 | 227.00 |
| Vermont | 1.00 | 0.00 | 1.00 | 0.00 | 0.00 | 0.00 |
| Virginia | 1852.00 | 0.00 | 1852.00 | 2211.00 | 0.00 | 2211.00 |
| Washington | 160.00 | 0.00 | 160.00 | 168.00 | 0.00 | 168.00 |
| West Virginia | 146.00 | 0.00 | 146.00 | 175.00 | 0.00 | 175.00 |
| Wisconsin | 451.00 | 0.00 | 451.00 | 545.00 | 0.00 | 545.00 |
| Wyoming | 58.00 | 0.00 | 58.00 | 68.00 | 0.00 | 68.00 |

Similarly, column “Total outflow” denotes the expected total outgoing ventilators from a state *n* ∈ *𝒩* to FEMA in order to be shared with other states to be used to treat COVID-19 patients, and is calculated as

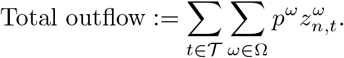

Also, column “Net flow” represents the difference between “Total inflow” and “Total outflow”.

The results in Table 4 indicates that in Cases VI (Severe) and V (Extreme), the State of New York requires between 11,100-17,500 additional ventilators for COVID-19 patients during its peak demand. However, between 400 to 17,000 of these ventilators can be given to a different state after the peak demand in the State of New York has subsided. The insights about other states can also be obtained from this table.

#### 4.5.2 The Effect of Early Ramp-up in Production on Ventilators’ Shortage

In this section, we consider the cases that the ramp-up in production happens on 04/01/2020 or 04/07/2020, as opposed to the baseline production, where the ramp-up in production happens on 04/15/2020. A summary of ventilators’ shortage is given in Table 5 for the parameter setting (*γ, ρ, τ*) = (0.75, 3.00, 0), under the two worst demand situations, Cases V (Severe) and VI (Extreme). As it is evident from Table 5, early ramp-up in production could save up more than 80,000 and 100,000 lives in Case V (Severe) and Case VI (Extreme), respectively.

**Table 5:**
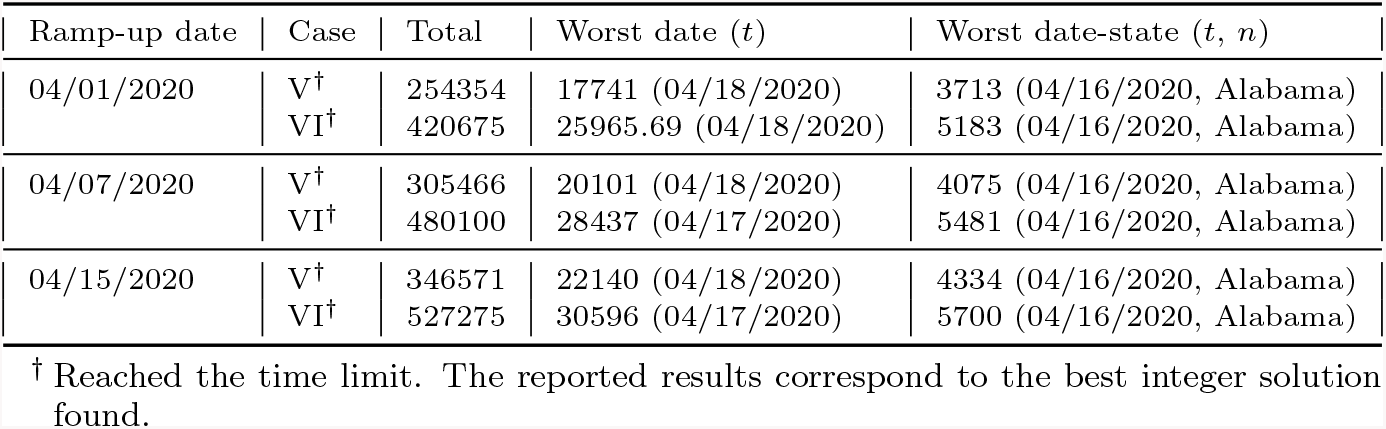
Ventilators’ shortage summary with early ramp-up in production under Cases V and VI, with *Q* = 320 and (*γ, ρ, τ, L*) = (0.75, 3.00, 0.00, 0).

| Ramp-up date | Case | Total | Worst date ($t$) | Worst date-state ($t, n$) |
| --- | --- | --- | --- | --- |
| 04/01/2020 | V<sup>†</sup> | 254354 | 17741 (04/18/2020) | 3713 (04/16/2020, Alabama) |
|  | VI<sup>†</sup> | 420675 | 25965.69 (04/18/2020) | 5183 (04/16/2020, Alabama) |
| 04/07/2020 | V<sup>†</sup> | 305466 | 20101 (04/18/2020) | 4075 (04/16/2020, Alabama) |
|  | VI<sup>†</sup> | 480100 | 28437 (04/17/2020) | 5481 (04/16/2020, Alabama) |
| 04/15/2020 | V<sup>†</sup> | 346571 | 22140 (04/18/2020) | 4334 (04/16/2020, Alabama) |
|  | VI<sup>†</sup> | 527275 | 30596 (04/17/2020) | 5700 (04/16/2020, Alabama) |

<sup>†</sup> Reached the time limit. The reported results correspond to the best integer solution found.

#### 4.5.3 The Effect of Lead-Time on Ventilators’ Shortage

In this section, we analyze ventilators’ shortage for the case that there is a lead-time of one day. A summary of results under Case VI is presented in Table 6. It can be seen from this table that, as expected, the inventory shortfall increases with an increase in the lead-time (approximately up to 500 on the worst day).

**Table 6:**
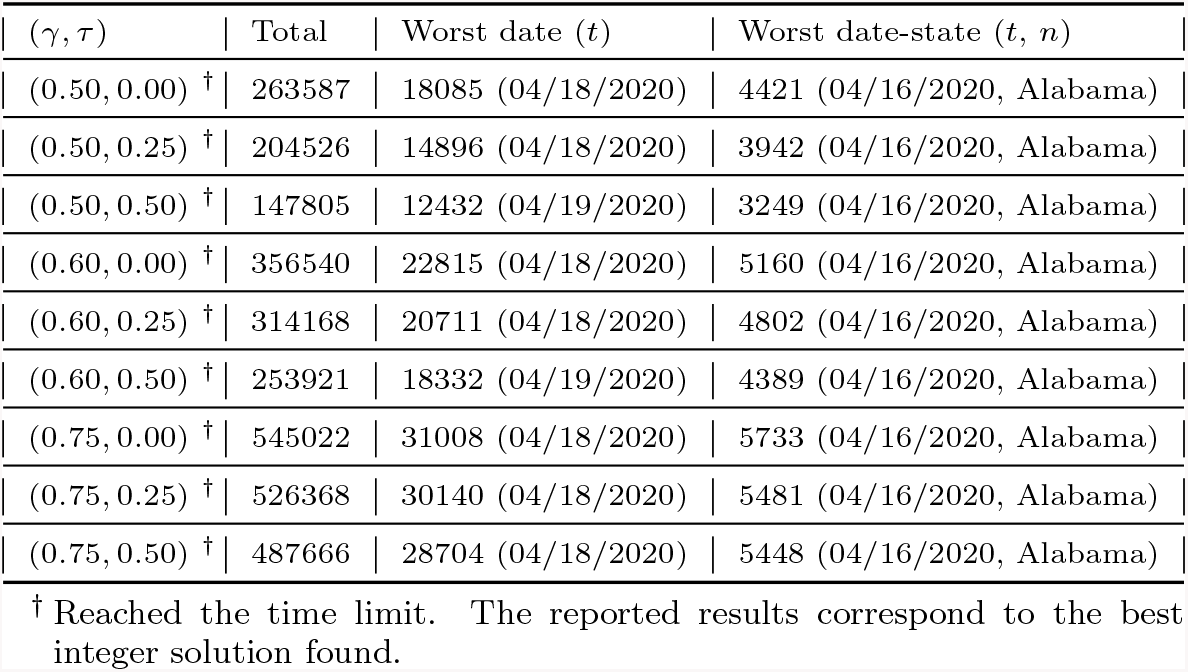
Ventilators’ shortage summary with a lead-time of one day, under Case V and VI, with *Q* = 320 and *ρ* = 3.00.

| $(\gamma, \tau)$ | Total | Worst date ($t$) | Worst date-state ($t, n$) |
| --- | --- | --- | --- |
| (0.50, 0.00) <sup>†</sup> | 263587 | 18085 (04/18/2020) | 4421 (04/16/2020, Alabama) |
| (0.50, 0.25) <sup>†</sup> | 204526 | 14896 (04/18/2020) | 3942 (04/16/2020, Alabama) |
| (0.50, 0.50) <sup>†</sup> | 147805 | 12432 (04/19/2020) | 3249 (04/16/2020, Alabama) |
| (0.60, 0.00) <sup>†</sup> | 356540 | 22815 (04/18/2020) | 5160 (04/16/2020, Alabama) |
| (0.60, 0.25) <sup>†</sup> | 314168 | 20711 (04/18/2020) | 4802 (04/16/2020, Alabama) |
| (0.60, 0.50) <sup>†</sup> | 253921 | 18332 (04/19/2020) | 4389 (04/16/2020, Alabama) |
| (0.75, 0.00) <sup>†</sup> | 545022 | 31008 (04/18/2020) | 5733 (04/16/2020, Alabama) |
| (0.75, 0.25) <sup>†</sup> | 526368 | 30140 (04/18/2020) | 5481 (04/16/2020, Alabama) |
| (0.75, 0.50) <sup>†</sup> | 487666 | 28704 (04/18/2020) | 5448 (04/16/2020, Alabama) |

<sup>†</sup> Reached the time limit. The reported results correspond to the best integer solution found.

## 5 Viewing Results using an Online Tool

A companion online planning tool is developed in order to view the outputs on the number of ventilators needed and the shortage that might happen under various conditions [9]. This website is available at https://covid-19.iems.northwestern.edu. The users can choose the demand scenario (Cases I–VI) and choose different options for parameter *γ*, the fraction of ventilators used to care for non-COVID patients, parameter *τ*, state’s willingness to share the fraction of their initial COVID-19-use ventilators, parameter *ρ*, the state’s risk-aversion to sending their idle ventilators to FEMA for use in a different state, and parameter *L* for lead-time. The results on the website are shown in interactive graphical and tabular formats. A snippet of this online planning tool is given in Figure 4.

**Figure 4:**
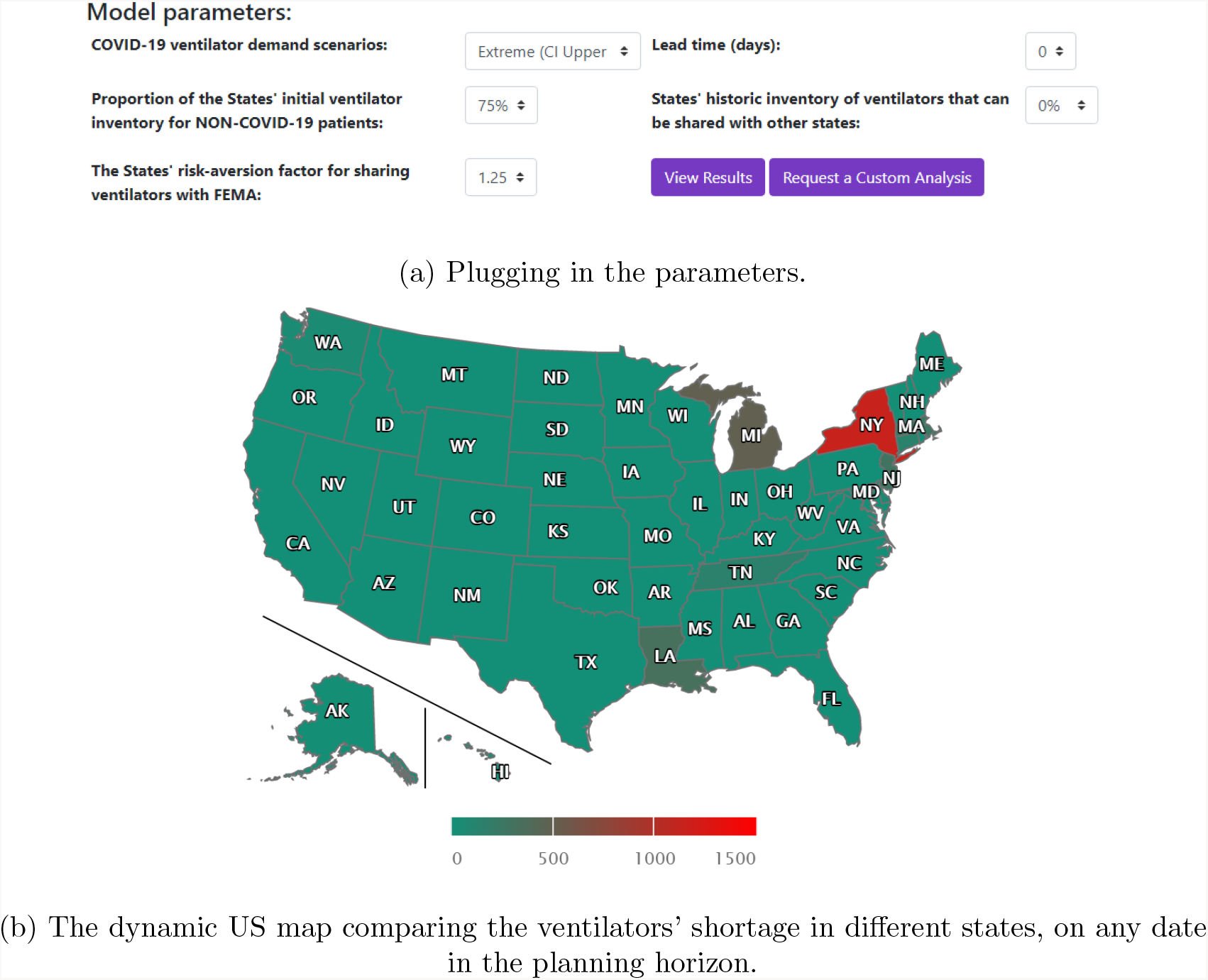
Online COVID-19 ventilator allocation and sharing planning tool [9].

Interested readers can refer to this online companion for more details and analysis beyond what is presented in this paper. The results on [9] will be updated as additional computations are conducted and new forecast confidence intervals become available.

## 6 Concluding Remarks

We have presented a model for procuring and sharing life-saving resources whose demand is stochastic. The demand arising from different entities (states) peaks at different times, and it is important to meet as much of this demand as possible to save lives. Each participating state is risk averse to sharing their excess inventory at any given time, and this risk-aversion is captured by using a safety threshold parameter. Specifically, the developed model is applicable to the current COVID-19 pandemic, where many US states are in dire need of mechanical ventilators to provide life-support to severely- and critically-ill patients. Computations were performed using realistic ventilator need forecasts and availability under a wide combination of parameter settings.

Our findings suggest that the fraction of currently available ventilators that are to be used for non-COVID-19 patients strongly impacts state and national ability to meet demand arising from COVID-19 patients. When more than 40% of the existing inventory is available for COVID-19 patients, the national stockpile is nearly sufficient to meet the demand in mild cases. However, if less than 25% of the existing inventory is available for COVID-19 patients, the current national stockpile and the anticipated production may not be sufficient under extreme demand scenarios. As expected, the magnitude of this shortfall increases when one considers more and more extreme demand scenarios.

Overall, the model developed in this paper can be used as a planning tool/framework by state and federal agencies in acquiring and allocating ventilators to meet national demand. The results reported in this paper can also provide a guide to states in planning for their ventilator needs. We, however, emphasize that these results are based on certain modeling assumptions. This include the process of demand forecast scenario generation, estimates of initial ventilator inventory, and future production quantities. Each one of these, as well as other model parameters, can be changed in the model input to obtain more refined results. Nevertheless, an important finding is that a state’s willingness to share its idle inventory can help address overall shortfall.

While this paper has focused on ventilator needs in the US, such a model can also be adapted for use in international supply-chain coordination of equipment such as ventilators across countries. COVID-19 is expected to have different peak dates and demand cycles in other countries, and one or two additional disease spread cycles are likely until an effective vaccine becomes available.

In conclusion, we point out that the model developed in this paper has a one-time planning decision, i.e., there are no “wait-and-see” decisions in the model over time. One can also formulate the ventilator allocation problem as a time-dynamic multistage stochastic program, where the decision maker can make recourse decisions as time evolves based on the information available so far on the stochastic demands and past decisions. We are currently working on such an extension.

## Data Availability

Availability of data used to conduct the research reported in this paper is either referenced in the paper, or it is available from the authors.

https://covid-19.iems.northwestern.edu/

## References

[1] State Health Systems Strained as Coronavirus Outbreak Spreads. https://www.cnbc.com/2020/03/29/state-health-systems-strained-as-coronavirus-outbreak-spreads.html [Last accessed on March 31, 2020].

[2] Mapping US Health System Capacity Needs to Care for COVID-19 Patients. https://www.covidcaremap.org [Last accessed on March 31, 2020].

[3] NY Governer Andrew Cuomo Holds Coronavirus Briefing-NBC News. https://www.youtube.com/watch?v=uIhRM9pnH40. [Last accessed on March 31, 2020].

[4] IHME COVID-19 Projections. https://covid19.healthdata.org/projections [Last accessed on April 02, 2020].

[5] COVID-19 Resource Center. https://coronavirus.jhu.edu/map.html [Last accessed on March 31, 2020].

[6] United States Resource Availability for COVID-19. https://sccm.org/Blog/March-2020/United-States-Resource-Availability-for-COVID-19 [Last accessed on March 31, 2020].

[7] Coronavirus Outbreak: Trump Invokes Defense Production Act (DPA),. https://www.youtube.com/watch?v=V9Il0D9o3io&t=195s [Last accessed on March 31, 2020].

[8] March 31 White House Briefing from Coronavirus Task Force,. https://www.youtube.com/watch?v=CWFOKznwNhQ [Last accessed on March 31, 2020].

[9] COVID-19 Ventilator Allocation and Sharing Planning Tool. https://covid-19.iems.northwestern.edu.

[10] O. M. Araz, A. Galvani, and L. A. Meyers. Geographic prioritization of distributing pandemic influenza vaccines. Health Care Management Science, 15:175–187, 2012.

[11] B. Armbruster and M. L. Brandeau. Contact tracing to control infectious disease: when enough is enough. Health Care Management Science, 10:341–355, 2007.

[12] D. Bienstock and A. C. Zenteno. Models for managing the impact of an epidemic, 2015. arXiv preprint 1507.08648 [math.OC].

[13] J. R. Birge and F. Louveaux. Introduction to Stochastic Programming. Springer, New York, 2nd edition, 2011.

[14] H. Cao and S. Huang. Principles of scarce medical resource allocation in natural disaster relief: a simulation approach. Medical Decision Making, 32(3):470–476, 2012.

[15] S. R. Earnshaw, K. Hicks, A. Richter, and A. Honeycut. A linear programming model for allocating HIV prevention funds with state agencies: a pilot study. Health Care Management Science, 10:239–252, 2007.

[16] Y. Feng, I. Wu, and T. Chen. Stochastic resource allocation in emergency departments with a multi-objective simulation optimization algorithm. Health Care Management Science, 20:55–75, 2017.

[17] S. Flessa. Where efficiency saves lives: A linear program for the optimal allocation of health care resources in developing countries. Health Care Management Science, 3:249–267, 2000.

[18] F. Gorunescu, S. I. McClean, and P. H. Millard. Using a queueing model to help plan bed allocation in a department of geriatric medicine. Health Care Management Science, 5:307–312, 2002.

[19] H.-C. Huang, O. M. Araz, D. P. Morton, G. P. Johnson, P. Damien, B. Clements, and L. A. Meyers. Stockpiling ventilators for influenza pandemics. Emerging Infectious Diseases, 23(6), 2017.

[20] M. Hui, Azhar. The continuing 2019-ncov epidemic threat of novel coronaviruses to global health—the latest 2019 novel coronavirus outbreak in wuhan,china. International Journal of Infectious Disease, 91(1):264–266, 2020.

[21] E. H. Kaplan. Containing 2019-ncov (Wuhan) coronavirus. Health Care Management Science, 23:1–4, 2020.

[22] M. I. Meltzer, A. Patel, A. Ajao, S. V. Nystrom, and L. M. Koonin. Estimates of the demand for mechanical ventilation in the united states during an influenza pandemic. Clinical Infectious Diseases, 60(Suppl 1:S52-7):88–94, 2015.

[23] L. Rosenbaum. Facing covid-19 in italy ethics, logistics, and therapeutics on the epidemic’s front line. New England Journal of Medicine, 2020.

[24] L. Rubinson, F. Vaughn, S. Nelson, S. Giordano, T. Kallstrom, T. Buckley, T. Burney, N. Hupert, R. Mutter, M. Handrigan, and et al. Mechanical ventilators in us acute care hospitals. Disaster Medicine and Public Health Preparedness, 4(3):199–206, 2010. doi: 10.1001/dmp.2010.18.

[25] I. C.-. H. service utilization forecasting team and C. J. Murray. Forecasting COVID-19 impact on hospital bed-days, ICU-days, ventilator days and deaths by US state in the next 4 months. MedRxiv, 2020. https://doi.org/10.1101/2020.03.27.20043752.

[26] A. Shapiro, D. Dentcheva, and A. Ruszczyński. Lectures on Stochastic Programming: Modeling and Theory. MPS-SIAM Series on Optimization. Society for Industrial and Applied Mathematics, Philadelphia, USA, 2nd edition, 2014.

[27] C. Vasilakis and E. EI-Darzi. A simulation study of the winter bed crisis. Health Care Management Science, 4:31–36, 2001.

[28] Z. Wu and J. M. McGoogan. Characteristics of and important lessons from the coronavirus disease 2019 (covid-19) outbreak in china: summary of a report of 72 314 cases from the chinese center for disease control and prevention. JAMA, 2020.

[29] Y. Xiang and J. Zhang. A medical resource allocation model for serving emergency victims with deteriorating health conditions. Annals of Operations Research, 236(1):177–196, 2016.

[30] S. Zaza, L. M. Koonin, A. Ajao, S. V. Nystrom, R. Branson, A. Patel, B. Bray, and M. F. Iademarco. A conceptual framework for allocation of federally stockpiled ventilators during large-scale public health emergencies. Health Security, 14(1):88–94, 2016.

